# Early transcriptional signature in chikungunya predicts chronic arthralgia and reveals deficient antiviral response

**DOI:** 10.64898/2026.05.04.26352362

**Authors:** Thiago Cerqueira-Silva, Laise de Moraes, Blenda de Jesus Pereira, Jessica Silva, Cibele Orge, Kevan Akrami, Ligia Souza, Laila Horta, Moacyr Rego, Amanda Albuquerque, Joyce Karoline da Silva, Paula Cassais, Luciano Pamplona Calvacante, Cristina Ribeiro Cardoso, Pablo Ivan P. Ramos, Luciane Amorim Santos, Manoel Barral-Netto, Aldina Barral, Ricardo Khouri, Viviane S. Boaventura

## Abstract

**Objective:** Long-term sequelae following viral infections, such as Chikungunya and SARS-CoV-2, are associated with persistent symptoms, with a notably higher prevalence in women. This study investigated the early determinants of progression to chronic chikungunya (CC) and examined the specific role of biological sex on disease outcomes.

**Methods:** We analysed peripheral blood mononuclear cells (PBMCs) sampled within seven days of disease onset, recruited between 2016 and 2020. The study compared patients who eventually recovered (RC, n = 11) with those who progressed to develop CC (n = 24). We analysed gene signatures through transcriptomics and validated the results using qRT-PCR and flow cytometry

**Results:** Ten genes were differentially expressed between the cohorts. Specifically, the study identified an upregulation of IKZF2 (encoding Helios) in CC patients, which was confirmed by qRT-PCR. Conversely, ACKR3 (encoding a CXCL12 scavenger in the ACKR3/CXCR4/CXCL12 axis) was upregulated in RC patients and validated by flow cytometry. Furthermore, CC cases demonstrated higher viral loads and downregulation of IFN-α and IFN-γ pathways. We also found that immune profiles differed between men and women; specifically, interferon α/γ and TNF signalling pathways were upregulated in women with CC but downregulated in men with CC relative to recovered individuals.

**Discussion:** Immune profiles differed significantly between men and women within both the CC and RC groups. These findings suggest that progression to chronic disease is influenced by an impaired early antiviral response combined with sex-specific immune regulation. Furthermore, *ACKR3* and *IKZF2* are identified as potential prognostic biomarkers for chronic chikungunya.

## Introduction

Post-acute viral syndrome (PAVS) refers to persistent symptoms and health issues lasting weeks to years after an acute infection. Long-term pain, myalgia, and fatigue have been reported following Chikungunya virus (CHIKV) or SARS-CoV-2 exposure.[1–3] Chikungunya, a vector-borne arthritogenic disease reported in over 100 countries, has emerged as a major public health problem in recent decades due to a rapid geographic spread and the high risk of PAVS.[4,5] A high proportion of individuals suffer from long-standing, debilitating disease. After CHIKV infection, up to 87% of patients develop a form of PAVS called chronic chikungunya (CC), characterised by joint pain persisting for over three months, among other symptoms.[6] Like other PAVS, women face a higher risk of developing CC,[7,8] and there is currently no specific treatment for the disabling consequences at long-term post-infection. Despite the recent approval of a live attenuated CHIKV vaccine,[9] which is contraindicated in certain populations, such as immunocompromised individuals, thus limiting the immunisation potential for broad population-level protection. This scenario offers a unique opportunity to investigate the mechanisms of PAVS and their impact on human health. Then, unravelling the processes driving CC may contribute to the development of targeted therapies and enhance our understanding of the immunopathogenesis of other PAVS.

Early identification of individuals likely to develop CC could optimise medical intervention and reduce the disease’s global impact. Some molecules, including *IL-12*, *IFN*-γ, *IL-6*, and *GM-CSF*, have been proposed as potential prognostic biomarkers for CC,[10,11] although validation is still lacking. OMICS approaches, which enable the simultaneous screening of thousands of molecules, have been successfully applied to various conditions, enhancing biomarker precision and uncovering pathways involved in disease mechanisms.[12]

In line with that, we conducted a comparative analysis of the viral and host transcriptional profiles from PBMCs isolated during the acute phase of CHIKV infection. We focused on distinguishing patients who achieved full recovery (RC) from those who developed CC, with persistent symptoms lasting over one year. Early biomarkers predictive of chronicity, identified via transcriptional profiling, were experimentally validated at mRNA and protein levels. Additionally, we explored sex-specific differences in transcriptional signatures, with a dedicated analysis comparing male patients who progressed to the CC versus the RC group. This dual approach, combining discovery, experimental validation, and sex-stratified analysis, provided insights into the molecular mechanisms underlying chronic CHIKV pathogenesis and potential diagnostic, prognostic, or therapeutic targets for this disabling disease.

## Methodology

### Study population

We randomly selected 35 (CC =24 and recovered=11) cases from a cohort of 134 chikungunya patients from three cities (Itabuna, Campo Formoso and Maranguape) between 2016 and 2018, as described previously.[8] Briefly, mono CHIKV infection was confirmed by a positive qRT-PCR in blood, saliva or urine samples, with negative qRT-PCR or IgM tests for Dengue or Zika viruses. We conducted structured questionnaires and medical examinations at recruitment (within 10 days of symptom onset) and at 3, 9, and 12 months after symptom onset. All data was stored using REDCap. We also enrolled nine healthy controls from the same area who tested negative for Chikungunya, Dengue, and Zika viruses. The flow cytometry experiments were conducted in patients recruited in El Salvador in 2020, following the same procedures as those used in the previous cohort.

### Ethics

This study was approved by the Institutional Review Board of the School of Medicine—Federal University of Bahia—Brazil (approval number: 1.657.324)

### RNA extraction from PBMCs

Blood samples were collected at the time of recruitment. Total RNA was extracted from peripheral blood mononuclear cells (PBMC) of CHIKV cases and healthy controls using PureLink RNA Mini Kit (Thermo Fisher Scientific) and treated with DNase I, Amplification Grade (Thermo Fisher Scientific). The concentration of the material was confirmed using Qubit 3 fluorometer (Thermo Fisher Scientific) with Qubit RNA HS Assay Kit (Thermo Fisher Scientific), and its integrity was assessed with a Bioanalyzer 2100 (Agilent) with RNA 6000 Nano Kit (Agilent). The depletion of the rRNA was performed using the Ribo-zero rRNA removal kit (Illumina).

### Library construction for RNA sequencing

The RNA-Seq library preparation was performed using TruSeq Stranded Total RNA Library Prep (Illumina) and sequenced using the NextSeq (Illumina) with a fragment size of 75bp. All procedures were conducted according to the manufacturer’s specifications.

### Virus detection

Unmapped reads were aligned to custom databases containing Chikungunya, Dengue, and Zika viruses (GenBank codes: AF369024.2, U88536.1, U87411.1, AY099336.1, AF326825.1, AY632535.2) using viGen bioinformatic pipeline.[39]

### cDNA synthesis and qRT-PCR

Total RNA was used to synthesis cDNA using SuperScript IV First-Strand Synthesis System (Thermo Fisher Scientific) with 50µM Oligo d(T)20, according to the manufacturer’s instructions. The RT-qPCR experiments were performed on a 7500 Real Time PCR System (Applied Biosystems) using TaqMan Universal PCR Master Mix (Thermo Fisher Scientific). The primers used were from the TaqMan Gene Expression Assay and included: IKZF2 (Hs00212361_m1), NAV3 (Hs00372108_m1), GAPDH (Hs02786624_s1), PTPRC (Hs04189704_m1), TMEM176A (Hs00962650_m1), TMEM176B (Hs00218506_m1), ACKR3 (Hs00664172_m1). All protocols were conducted following the manufacturer’s instructions. In addition to GAPDH, we also included the PTPRC (CD45) gene as a reference to stabilise gene expression in relation to leukocyte cells.[40] Samples with unreliable values in the qRT-PCR, i.e. Ct values higher than 35, were excluded from the analysis.[41]

### Data Analysis

The raw sequence files generated underwent quality control analysis using *FastQC v0.12.1*.[42] After visual inspection of quality results, the samples were aligned to the human reference genome using *STAR 2.7.11a,[43] samtools v1.18*,[44] *ribodetector v0.2.7[45]* was used to detect and remove residual rRNA sequences. Gene-level counts from RNAseq were quantified using *Subread featureCounts v2.0.6*,[46] reads overlapping features (exons) were counted for each feature. After removing low-count genes (less than 1 fragment per million in less than 10 samples), the raw counts were imported to R/Bioconductor package DESeq2 *v1.40.2*[47] to identify genes differentially expressed among samples. Genes were considered differentially expressed for adjusted p values < 0.05 and absolute log2 fold change > 2 in the pair infected-healthy analysis, and adjusted p values < 0.1 and absolute log2 fold change ≥1 in intra-group CC versus recovered. All comparisons were adjusted for sex. We additionally repeated the analysis stratifying by sex.

Gene set enrichment analysis (GSEA)[48] was applied using the package *fgsea v1.26.0*.[49] We used the pre-ranked mode, and an ordered gene list was produced per dataset by scoring each gene according to the following formula:

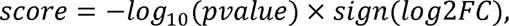

where *p*value is the unadjusted p-value from the differential expression analysis for a given gene, and log2FC is the corresponding log2 fold-change value derived from comparisons between groups. Similarities in gene dysregulation patterns between groups were evaluated by the Normalized Enrichment Score. We used the MSigDB v2023.2.Hs (Oct 2023) gene-sets available from the Molecular Signature Database. This database is a curated collection of 50 well-defined biological processes, providing refined and concise inputs for gene set enrichment analysis.[50] The difference in the viral copy and ΔΔCt was analysed using the Wilcoxon rank sum test.

### Luminex

Cytokine and soluble receptor quantification: Cytokine and soluble receptor levels were measured in plasma using Luminex-based multiplex assays (Merck Millipore). The premixed MILLIPLEX MAP Human Cytokine/Chemokine Magnetic Bead Panel was used to quantify EGF, G-CSF, GM-CSF, IFN-α2, IFN-γ, IL-1α, IL-1β, IL-1RA, IL-2, IL-3, IL-4, IL-5, IL-6, IL-7, IL-8, IL-10, IL-12(p40), IL-12(p70), IL-13, IL-15, IL-17A, IP-10, MCP-1, MIP-1α, MIP-1β, TNF-α, TNF-β, VEGF, and Eotaxin/CCL11. The MILLIPLEX Human Soluble Cytokine Receptor Panel was used to quantify sCD30, sEGFR, sgp130, sIL-1RI, sIL-1RII, sIL-2Rα, sIL-4R, sIL-6R, sRAGE, sTNFRI, sTNFRII, sVEGFR1, sVEGFR2, and sVEGFR3. The MILLIPLEX TGF-β1 Magnetic Bead Single Plex Kit was used to measure TGF-β1, and a custom-made MILLIPLEX Magnetic Bead Single Plex Kit was used to measure SDF-1α+β. All assays were carried out according to the manufacturer’s instructions. Acquisition was performed on a Luminex® 200 System (Luminex Corp), and median fluorescence intensity (MFI) values were obtained for each analyte.

### Flow Cytometry

Peripheral blood samples were collected at the time of recruitment, and PBMCs were isolated and cryopreserved. Prior to flow cytometry analysis, cells were thawed, and 1.5 × 10⁶ cells were used for staining of T cells and monocytes. To identify live cells, samples were incubated with the LIVE/DEAD™ Yellow Fixable Kit (Thermo Fisher) at a 1:1000 dilution in 1X PBS for 15 minutes at room temperature in the dark. After viability staining, monoclonal anti-human antibodies targeting surface markers were added and incubated for 30 minutes at 4°C in the dark. The following antibodies were used: CD45 PE-Cy5 [clone HI30] (BioLegend); CD4 BV650 [clone SK3] (BD Biosciences); CD8 APC-H7 [clone SK1] (BD Biosciences); PD-1 Alexa 700 [clone EH12-2H7] (BioLegend); CD16 PE [clone 3G8] (BD Biosciences); CD14 V500 [clone M5E2] (BD Biosciences); CXCR4 BV711 [clone L276F12] (BioLegend); and CXCR7 [clone 10D1] (BD Biosciences). For intracellular staining, the Transcription Factor Buffer Set (BD Biosciences) was used. Cells were fixed and permeabilised with 1X Perm/Wash buffer for 20 minutes at 4°C in the dark, followed by overnight incubation at 4°C on a shaker in 50 µL of BD Perm/Wash buffer containing fluorochrome-conjugated antibodies. The intracellular markers included FoxP3 PE-CF594 [clone 236A/E7] (BD Biosciences) and Helios [clone 22F6] (BioLegend). To detect the Chikungunya virus (CHIKV), a monoclonal anti-CHIKV antibody [clone CHK-166] (Thermo Fisher) was conjugated to Alexa Fluor 488 using the Zenon Kit (Thermo Fisher) and incubated for 1 hour at 4°C in the dark. Before acquisition, all samples were filtered through a 40-µm strainer. Data acquisition was performed using a BD Symphony™ flow cytometer (BD Biosciences), and data were analysed with FlowJo software (BD Biosciences). Considering the very low sample size, the difference in protein expression was analysed using T-test.[51]

## Results

### Early transcriptomic signatures of acute CHIKV infection reveal immune dysregulation associated with chronic disease progression

We first performed bulk RNAseq of PBMC samples from 35 patients during the acute phase of CHIKV infection. All patients were evaluated at least three months after symptom onset to assess the presence of arthralgia; among them, 24 developed chronic chikungunya (CC), and 11 fully recovered (RC). Cases were recruited a median of one day after symptom onset, and infection was confirmed by RT-PCR. Arthralgia in the CC group persisted for up to 949 days (median = 723, IQR = 380-949), whereas in the recovered group, the median duration was 11 days (IQR = 7-19). Both the CC and RC groups reported similar symptoms during the acute phase, except for edema, which was significantly more frequent among chronic cases (75% vs. 9%) (Table 1).

**Table 1:**
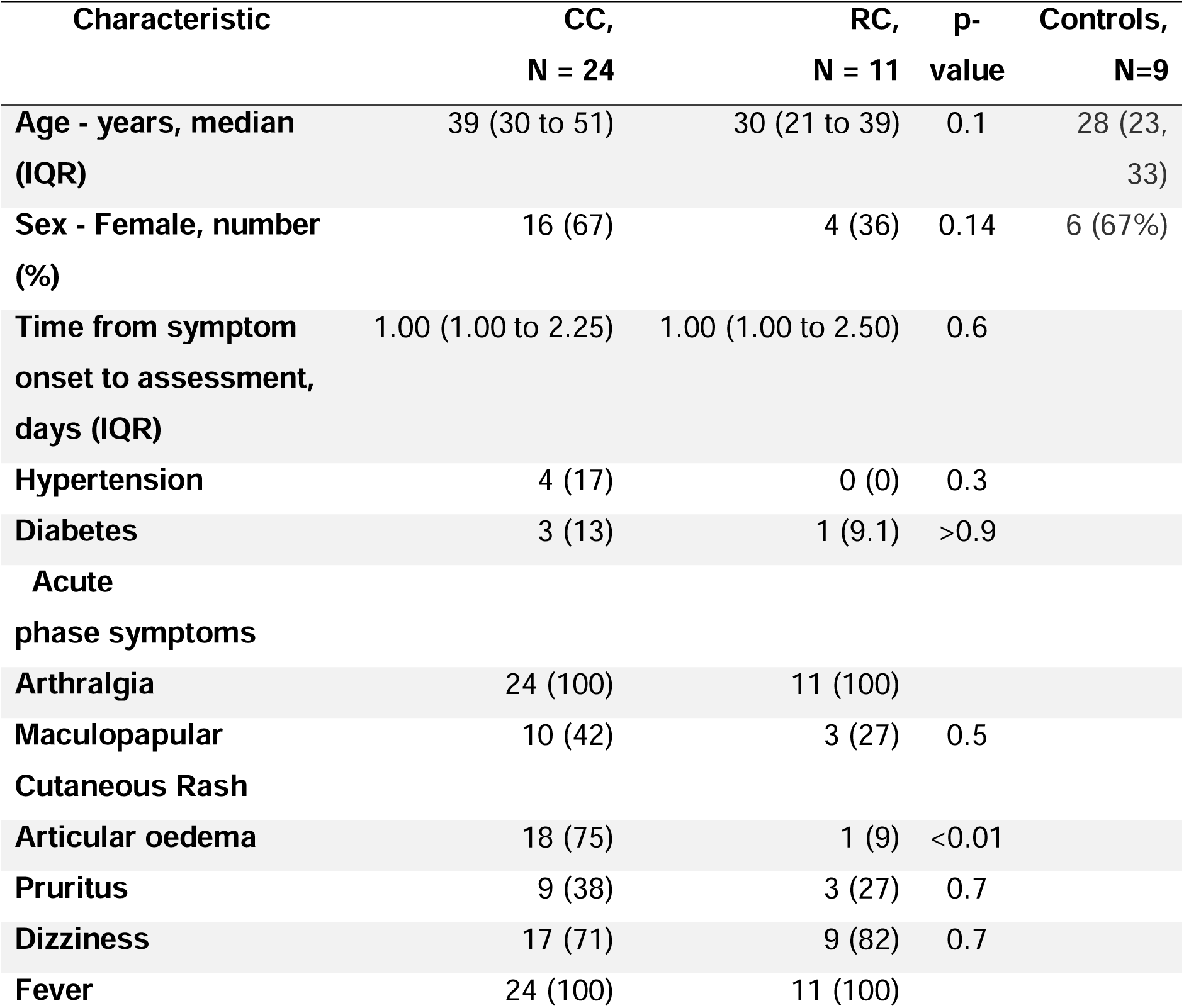
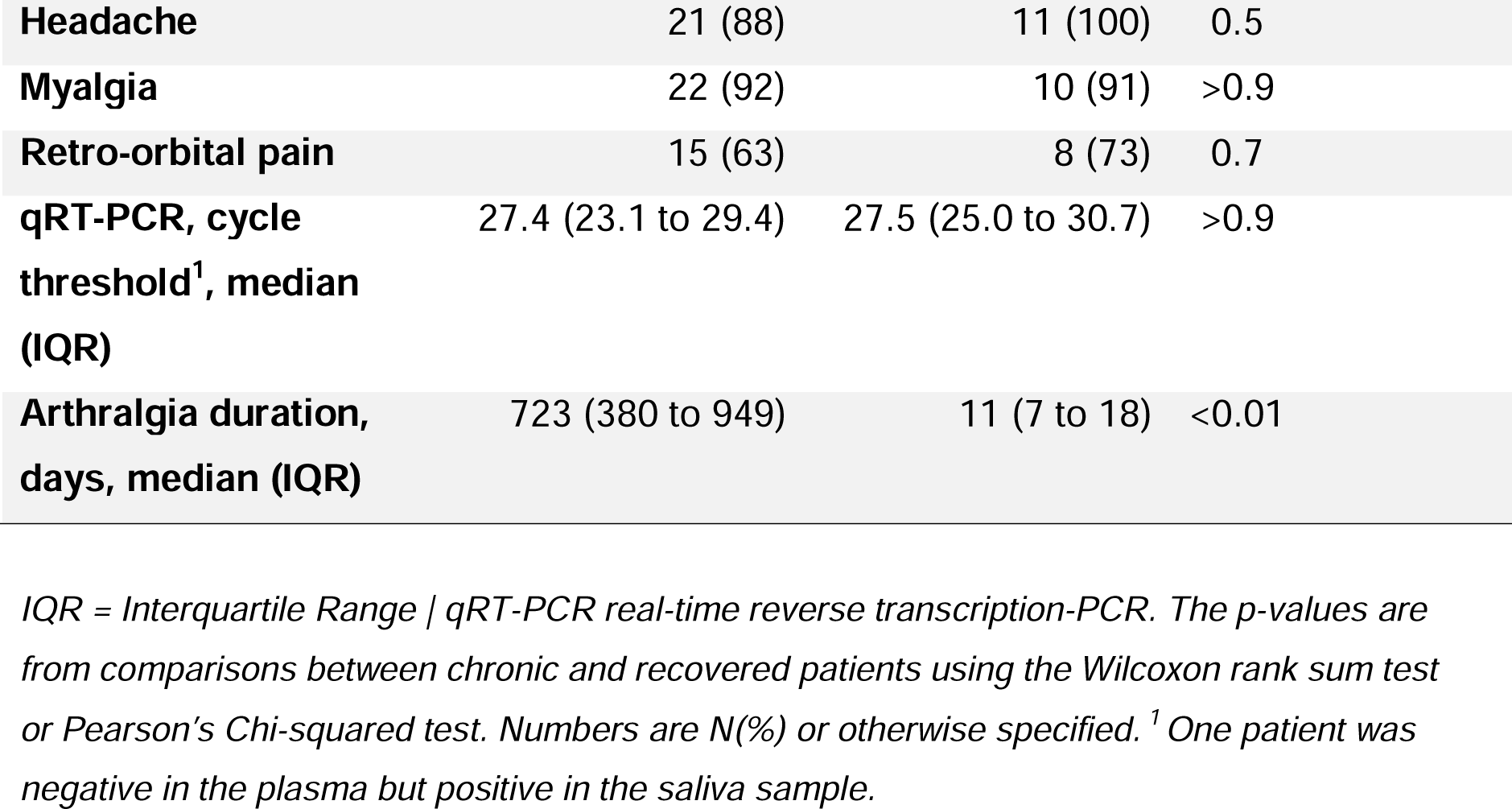
Baseline characteristics of patients with chronic chikungunya (CC), recovered (RC) individuals, and healthy controls.

The RNA-Seq data showed low contamination with ribosomal RNA (rRNA). In 40/44 samples, 98.2% ± 2.7% of the reads were retained in the pipeline. Four control samples had 47 to 69% of the reads retained after removing residual rRNA. After rRNA removal, our samples yield 86.6% ± 4.0 of mapping to the genome, with 79.8% ± 6.2 uniquely mapped.

We first compared the gene expression profile of CC and RC patients with that of healthy controls. We identified 1,240 differentially expressed genes (DEGs) in the CC group and 1,858 in the RC group (Supplementary files 1-3). Gene set enrichment analysis (GSEA) revealed a similar immune signature. Acute CHIKV infection activates several pathways associated with a strong pro-inflammatory response, with the most enriched signatures related to inflammation, coagulation, and type I (*IFN-α*) and II interferon responses (*IFN-γ*). However, some specific pathways exhibited higher negative enrichment scores in the CC group compared to the RC group, including *TNF*-signalling, *NF-κB* activation, *IFN-α* and *IFN-γ* responses, as well as hypoxia, adipogenesis, and protein secretion (Figure 1).

**Figure 1:**
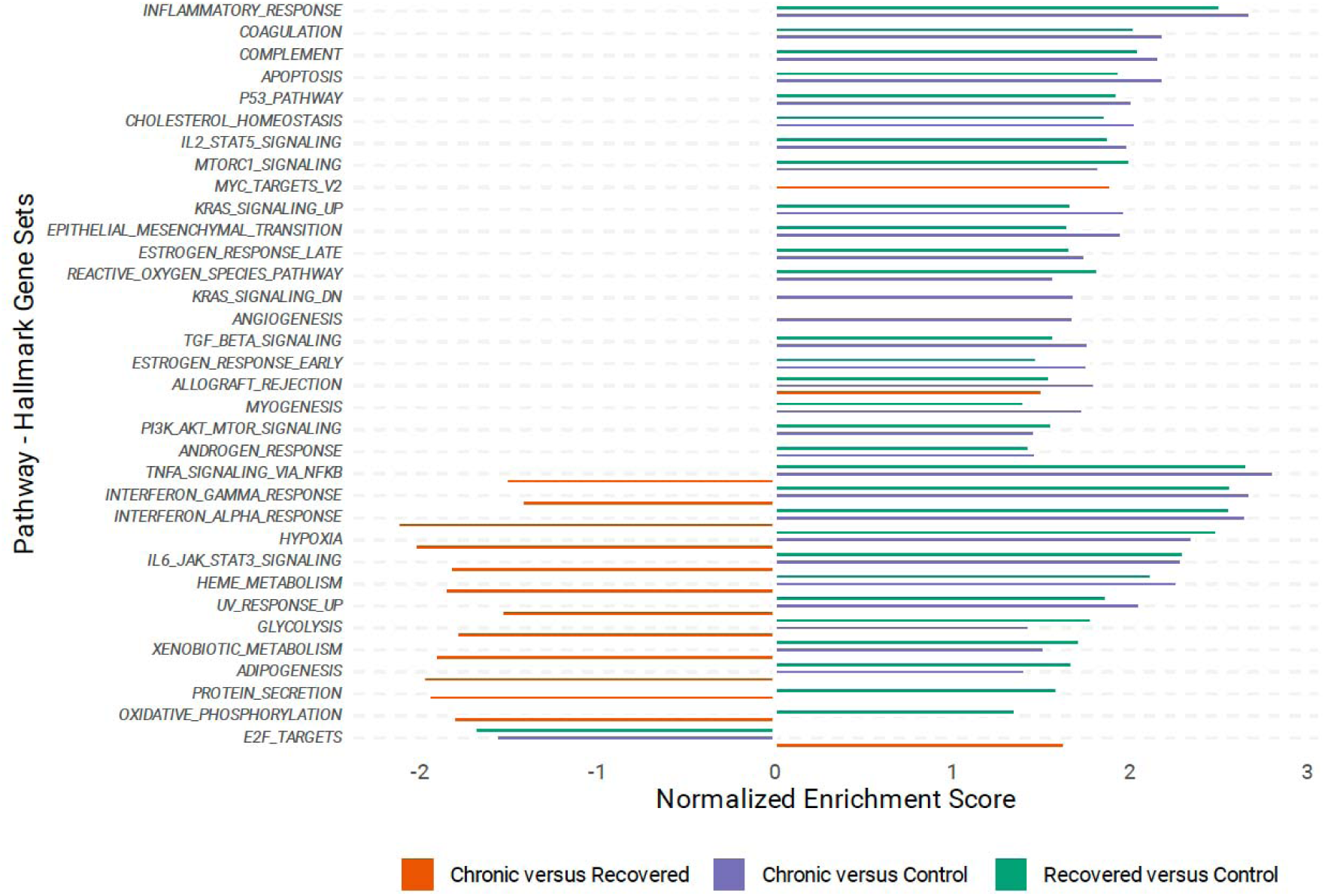
Gene set enrichment analysis results of Hallmark pathways. Only pathways with at least one comparison with a false discovery rate < 0.05 are exhibited.

We next compared DEGs between the CC and RC groups. Given the overall similarity in immune signatures between these groups, we applied less stringent thresholds for DEG identification (log₂ fold change ≥ 1 instead of log_2_ fold change>2, keeping FDR <0.05). Only ten genes met these criteria, four of which were significantly downregulated in CC, such as *ACKR3*, *TMEM176A*, *TMEM176B*, and *OLIG1* (Supplementary Figure 1). Notably, compared to healthy controls, CHIKV infection positively regulated the expression of *ACKR3* in RC but not in CC (Figure 2 and Supplementary Table 4). Six genes were significantly upregulated in CC compared to RC, including *IKZF2*, *NCS1*, *PTGER3*, *ST8SIA1*, *NAV3*, and *TNFRSF18* (Figure 2; Supplementary Table 3). CHIKV infection was associated with reduced expression of *IKZF2*, although the reduction was higher for the RC compared to the CC. In parallel, individuals in the CC group exhibited significantly higher levels of Chikungunya virus RNA in the RNA-Seq unmapped reads when compared to the RC group. The median number of viral copies was 12.6 (IQR: 2.8-31.1) in CC and 1.9 (1.0-2.8) in RC, respectively. (Figure 3A).

**Figure 2:**
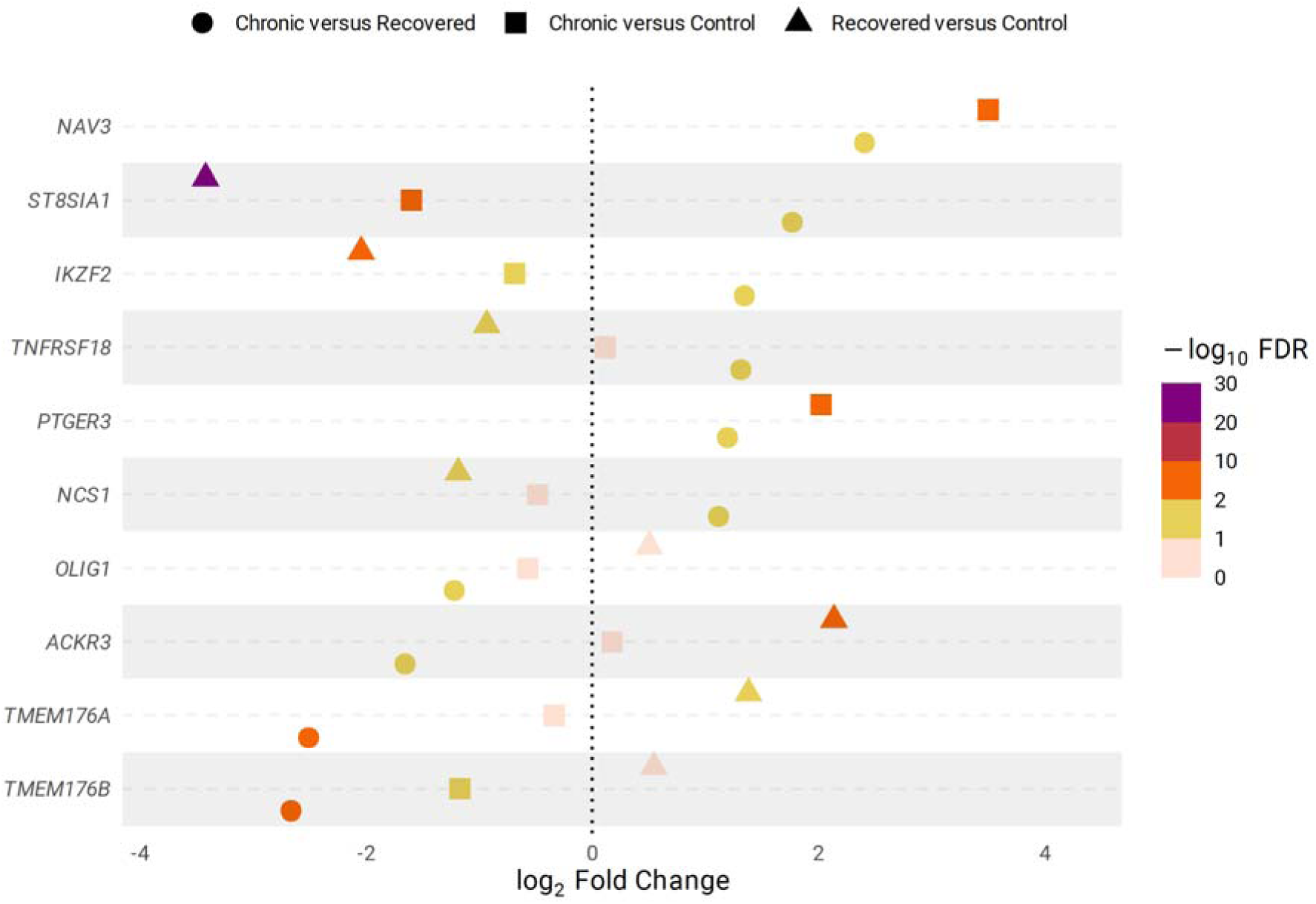
Dot plot showing log_2_ fold change and false discovery rate (FDR) of differentially expressed genes in chronic chikungunya, recovered. The comparison recovered versus control for NAV3 and PTGER3 was not conducted due to low counts (less than 1 fragment per million in less than 10 samples).

**Figure 3:**
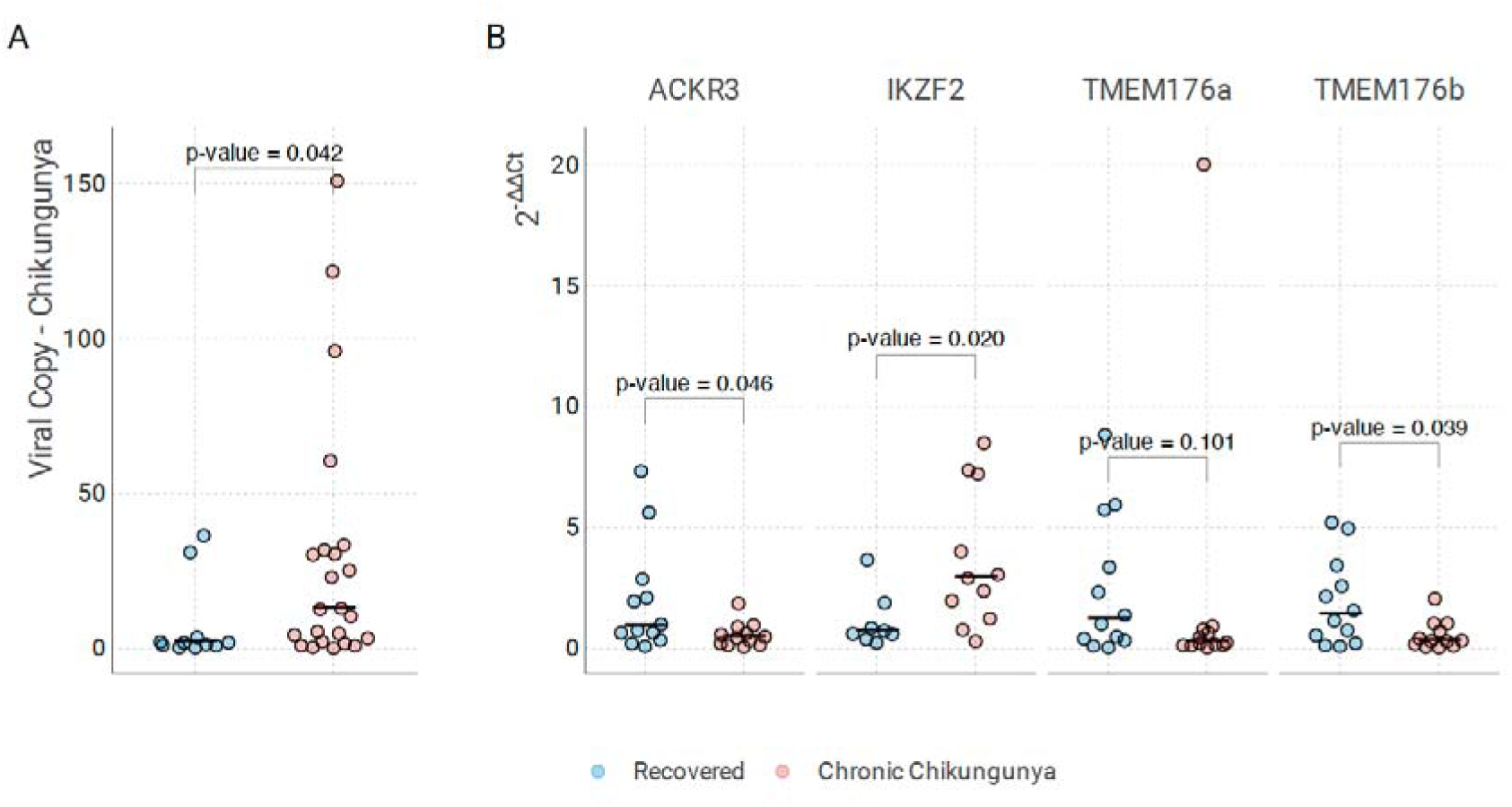
Dot plot showing: A) the number of viral copies detected in the unmapped reads of RNA-Seq; One sample was excluded due to no quantification of viral copies. B) the relative expression of selected genes using the 2^-ΔΔCt^ method (normalised by CD45 and GAPDH). The Black lines represent the median value in each group. Wilcoxon rank sum p-values.

### qRT-PCR validation identifies IKZF2 as a prognostic biomarker candidate for CC

To assess whether candidate genes identified by RNA-Seq could serve as prognosis biomarkers during acute CHIKV infection, we performed qRT-PCR analysis on PBMC samples from 24 patients (12 CC and 12 RC – Supplementary Table 5). We quantified the expression of *ACKR3*, *IKZF2*, *TMEM176a*, and *TMEM176b* (Supplementary Table 6). All four genes showed fold changes consistent with the RNA-Seq analysis: *ACKR3* (log_2_FC = -1.37, p-value 0.046), *IKZF2* (log_2_FC = 1.65, p-value 0.020), *TMEM176a* (log_2_FC = -1.52, p-value 0.101), and *TMEM176b* (log_2_FC = -1.69, p-value 0.039). (Figure 3B and Supplementary Table 5). *IKZF2* also showed significant results when normalising only by *GAPDH* (log_2_FC = 2.18, p-value 0.041). (Supplementary Table 6)

### Flow cytometry reveals selective upregulation of ACKR3 in the intermediate monocytes from recovered individuals

To validate the RNA-Seq results, we analysed PBMCs from a new group of seven patients recruited at a median of 4.5 days after symptom onset, all confirmed by RT-PCR. Four patients developed CC, and three recovered without persistent symptoms. Clinical demographics characteristics of patients were comparable between the CC and RC groups (Supplementary Table 7).

The *ACKR3* gene encodes *ACKR3*, a chemokine receptor for *CXCL12* that also interacts with *CXCR4*. *IKZF2* encodes *Helios*, a transcription factor with regulatory functions. We therefore examined *CXCR7* and *CXCR4* expression in classical (*CD14⁺CD16^-^*), intermediate (*CD14⁺CD16⁺*), and non-classical (*CD14^-^CD16⁺*) monocytes, as well as Helios expression in *CD4⁺FoxP3⁺* Tregs.

Recovered individuals showed higher *ACKR3* expression in intermediate monocytes compared to the chronic group, but similar levels in classical or non-classical monocytes (Figure 4A). Likewise, the *CXCR4* expression in all monocyte subsets (Figure 4A) and *Helios* expression in *CD4⁺* Tregs were similar in CC and recovered patients. (Figure 4 B/C).

**Figure 4:**
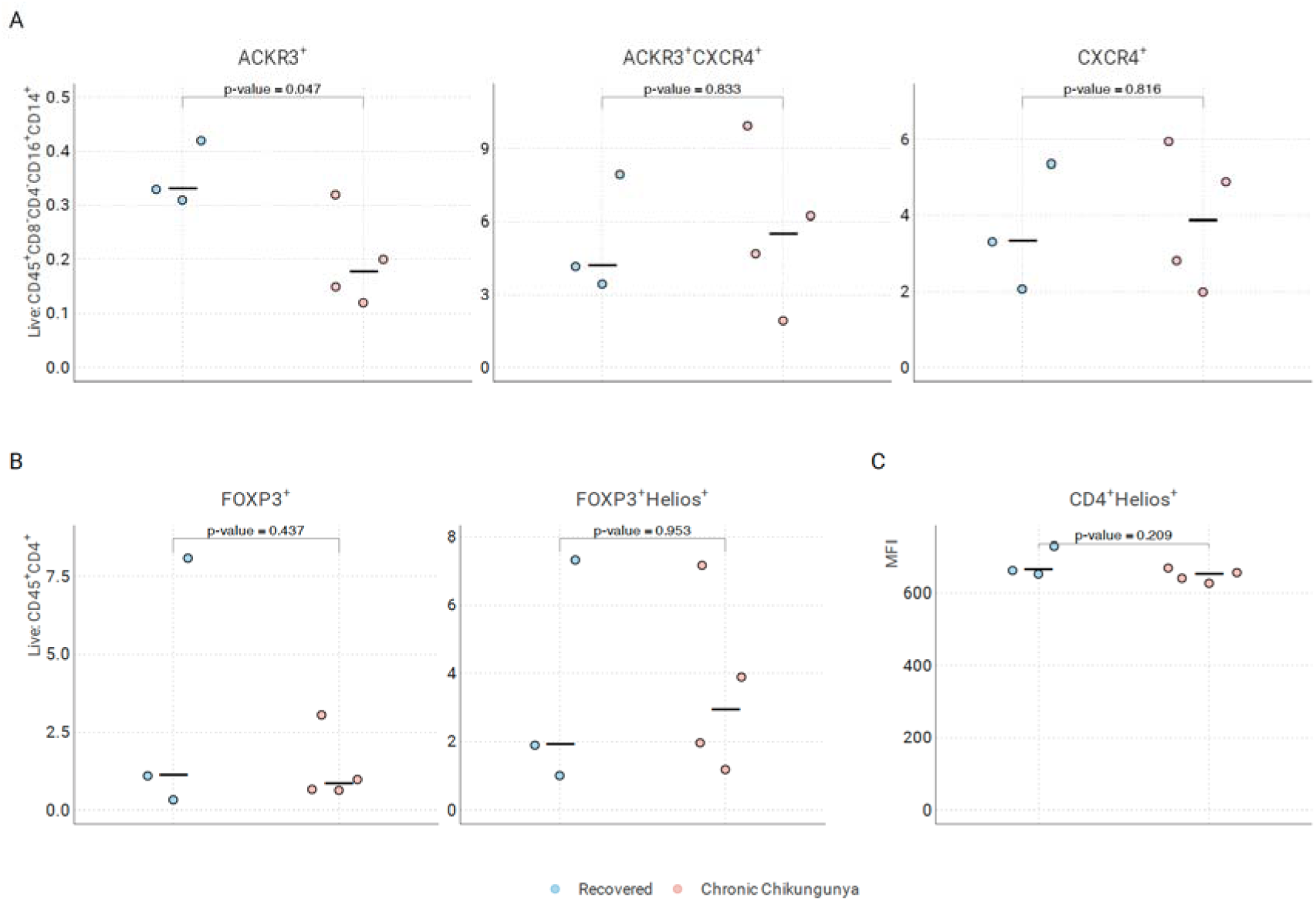
Flow cytometry analysis of intermediate monocytes (CD14⁺CD16⁺CXCR7⁺; CD14⁺CD16⁺CXCR7⁺CXCR4⁺; CD14⁺CD16⁺CXCR4⁺) and Tregs (CD4⁺FoxP3⁺; CD4⁺FoxP3⁺Helios⁺; CD4⁺Helios⁺) in recovered and chronic CHIKV individuals. Black lines indicate group medians. Gating schemes detailed in Supplementary Figure 2. P-values were calculated using a T-test.

**Figure 5.**
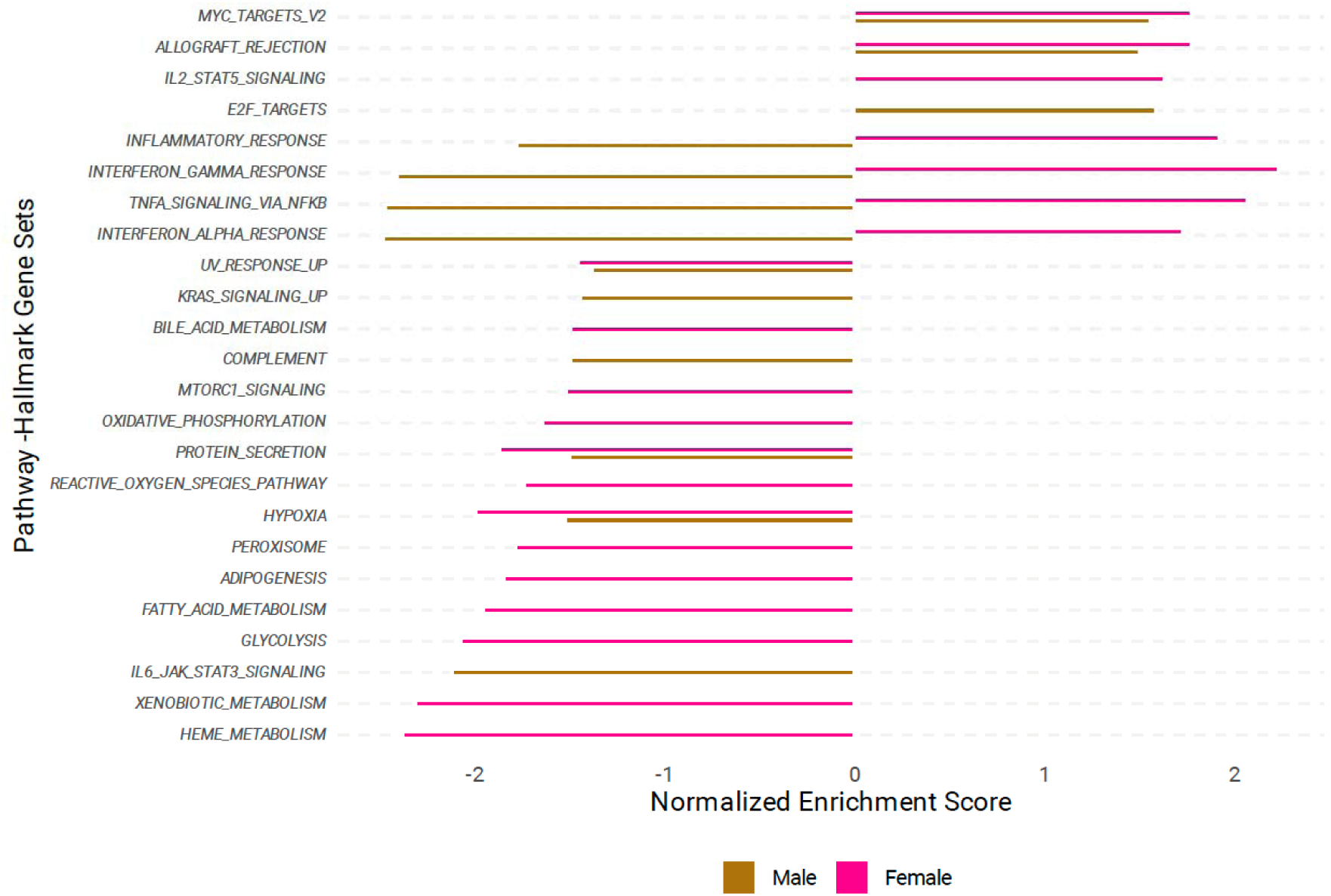
Gene set enrichment analysis results for Hallmark pathways comparing the chronic and recovered groups within each sex. Only pathways with at least one comparison with a false discovery rate < 0.05 are exhibited.

### Sex-Dependent Transcriptomic Signatures Associated with CC Progression

Given the higher prevalence and severity of CC clinical presentation in women, we next assessed whether distinct molecular signatures underlie the differential clinical outcomes between sexes and distinguish individuals who progress to chronic disease following acute CHIKV infection. In males, transcriptomic analysis revealed 145 DEGs, including 127 upregulated and 18 downregulated transcripts. (Supplementary Figure 3) Otherwise, in females, only a single differentially expressed gene (*VSIG4*) was identified, which was not differentially expressed in males. (Supplementary Figure 4) GSEA analysis revealed that only five gene sets were significantly enriched in the same direction across both sexes. Notably, pathways related to the inflammatory response displayed sex-specific regulation. In females, interferon *α/γ* and *TNF* signalling pathways were upregulated in CC compared to RC individuals, whereas in males, these pathways were downregulated in CC relative to RC. These findings indicate a sexual dimorphism in the transcriptional response to CHIKV infection, particularly in the transition from acute to chronic disease.

### Analysis of prognosis biomarkers in plasma

Lastly, we evaluated whether previously described biomarkers were predictors of CC in our cohort. We analysed 27 samples from the CC group and 11 from the recovered group, which were collected at a median of 1(IQR:1-3) day after symptom onset. (Supplementary Table 8) During acute infection, the expression of cytokines and chemokines in patients with CC and RC did not differ significantly (Supplementary Figure 5 and Supplementary Table 9).

## Discussion

This study identified a molecular signature observed in the first days of chikungunya onset associated with disease outcome. The genes differentially expressed in CC and RC are involved in regulatory activity, expression of pro-inflammatory and antiviral cytokines, cell migration, activation of patrolling anti-viral monocytes, and pain. Additionally, we detected sex-specific differences, suggesting distinct mechanisms associated with PAVS in chikungunya. These findings provide insight into the immunopathological mechanisms underlying PAVS after CHIKV infection.

We observed that, compared to RC, CC exhibited higher expression of *IKZF2*, a gene expressed mainly by *CD4^+^CD25^+^* regulatory T cells (Treg) and NK cells. [13–18] The ability to be a Treg depends on the expression of the FOXP3 gene[18]. The *FoxP3^+^Helios^+^* Tregs possess a high functional suppressive capacity and express *CXCR3* and *CCR4*, which allows transmigration to inflamed tissues.[19] Recent studies have shown a correlation between circulating *FoxP3^+^Helios^+^* cells and disease activity in patients with systemic lupus erythematosus.[20,21] The lower expression of *IKZF2* observed in recovered and CC groups compared to HC may be associated with a reduction in *FoxP3^+^Helios^+^* Treg subset, with a decreased immunosuppressive mechanism. In the first days of disease, reducing regulatory function may be important for developing an effective antiviral response. While lymphocytes represent the majority of PBMC in healthy and infected individuals, lymphopenia with an increased proportion of NK cells is commonly observed in acute CHIKV infection,[22] potentially increasing the representativeness of NK cells genes in the bulk analysis. The cell type responsible for *IKZF2* expression in PBMC of CC patients remains to be determined. Since these patients were recruited within three days of symptom onset, other investigations are required to assess the prognostic value of *IKZF2* at later time points.

Higher viremia during acute disease has been linked to CC.[23,24] In addition, the virus can spread and persist in the joint, causing inflammation and chronic pain in humans and non-human primates (NHP).[23,25] CHIKV has been detected in the joint of a patient suffering from chronic arthralgia years after disease onset.[23] In NHP, monocytes infected by CHIKV migrate to the joint early after infection.[25] By unveiling early events associated with prognosis, our findings support the hypothesis that PAVS after chikungunya is linked to virus persistence in target tissues. Investigating the molecular bases may help to elucidate the pathogenesis of other PAVS.

We observed relatively low expression of *ACKR3* in the CC group compared to the recovered group in both RNASeq and qRT-PCR. This chemokine, also known as *CXCR7*, composes an axis with *CXCR4* and its common ligand, *CXCL12*. *ACKR3* can modulate the axis downregulating *CXCL12* and *CXCR4* by scavenging or forming heterodimers, respectively.[26,27]. *CXCL12* complexed with *ACKR3* induces cell migration and a pro-inflammatory phenotype in leukocytes,[28] while *CXCL12* complexes with *CXCR4* promote cell migration to the synovium and induction of regulatory T cell activity.[13,29] Overexpression of *CXCR4* can be induced by CHIKV infection and seems to represent a viral escape mechanism.[30] Therefore, reduced *ACKR3* expression in CC may contribute to preferential activation of *CXCR4*, which may lead to a local downregulation of the antiviral response. The activation of the *CXCR4*/*CXCL12* axis has also been implicated in the development and maintenance of articular inflammation,[31] a hallmark of CC. On the other hand, recovered individuals exhibited higher levels of *ACKR3* compared to both CC and HC groups, potentially leading to reduced activation of the *CXCR4*/*CXCL12* axis. This downregulation of the *CXCR4*/*CXCL12* axis may limit the migration of regulatory T cells and other infected cells to inflamed tissues, thereby preventing viral dissemination to the synovium and subsequent chronic inflammation. Our finding of reduced expression of *ACKR3* in intermediate monocytes in the CC group compared to the RC also corroborates this hypothesis, as CHIKV stimulates the expansion of intermediate monocytes during acute chikungunya. The lower expression of *ACKR3* shifts the response towards an anti-inflammatory response and facilitates cell migration.[32,33] Taken together, these findings suggest that disruption of the *CXCR4/CXCL12* axis may be a protective mechanism in individuals who recover from CHIKV infection without progressing to chronic disease.

Other genes differentially expressed in CC and recovered have also been implicated in modulating the immune response. Compared to HC, we observed lower expression of *TNFRSF18* and *PTGER3* in RC patients. *PTGER3* mediates the anti-inflammatory effects of PGE2, and activation of *PTGER3* can inhibit pro-inflammatory cytokines, such as *TNF-α*, *IL-1β*, and *IL-6*.[34] *TNFRSF18* is highly expressed in Treg and can be considered a marker of Treg activity.[35] These data support the hypothesis that individuals who have fully recovered exhibit an immune response toward a pro-inflammatory milieu, which may be important in limiting virus activity during acute disease. Among the other differentially expressed genes comparing CC and recovered, *TMEM176 A/B*, and *NAV3* are also reported to present immunological functions. *TMEM176 A/B* were linked to efficient priming of naive *CD4*^+^ T cells by dendritic cells, whereas the *NAV3* gene has been linked to a shift from Th1-type to Th2-type in Sézary syndrome.[36,37] In our analysis, *TMEM176 A/B* genes showed higher expression in recovered, while *NAV3* was higher in chronic, which reinforcing the hypothesis of a weaker cellular response against the virus in CC, despite the known CHIKV epitopes recognized by CD4 T cells from patients presenting the chronic post viral arthralgia.[38] Finally, we could not find any previous articles indicating direct immunological functions for three DEGs (*OLIG1*, *ST8SIA1* and *NCS1*). Further studies should investigate the *in vitro* and *in vivo* activities of these DEGs.

The limited number of DEGs in females, in contrast to the broader transcriptional changes observed in males, suggests distinct regulatory mechanisms underlying chronic progression. The sex-specific modulation of key inflammatory pathways, upregulated in females and downregulated in males with CC, may reflect divergent immune trajectories that influence disease resolution versus persistence. Upregulation of *IFN* and *TNF-α* signalling in chronic female cases may indicate a sustained inflammatory state, potentially contributing to prolonged symptomatology. In contrast, downregulation of these pathways in males may point to insufficient immune activation or early resolution. These data underscore the importance of considering sex as a biological variable in the pathogenesis of post-viral syndromes and may inform the development of tailored therapeutic strategies.

Our study presents some limitations. First, we used a bulk transcriptome of PBMC, which does not allow us to determine the specific cell type that expresses the identified DEGs. While lymphocytes represent the majority of PBMC in healthy and infected individuals, lymphopenia with an increased proportion of NK cells is commonly observed in acute CHIKV infection [29], potentially increasing the representativeness of NK cells genes in the bulk analysis. Second, samples were collected approximately one day after symptom onset, when early innate immune cells, like NK cells, are more likely to be activated. This could bias the results to more accentuated differences in innate immune response than adaptive. Third, we could only evaluate four of the 10 DEGs detected in the RNA-Seq in a small subset of CHIKV patients from the cohort in the qRT-PCR experiments, which limited the validation step of this analysis. Fourth, in our analyses stratified by sex, the female group had a small number of recovered patients (four patients RC and 16 patients RC), reducing the power to detect differences in gene expression.

Our findings suggest that *ACKR3* and *IKZF2* can act as prognostic biomarkers for chronic chikungunya and that the early response involved in virus control during acute infection is related to disease progression. The predictive signature for CC varies by sex and suggests that persistence is associated with inefficient control of CHIKV infection and high viral loads. More research is needed to determine the role of these molecules in the anti-CHIKV response, as well as their involvement in PAVS.

## Data Availability

All data produced in the present work are contained in the manuscript. Sequence data that support the findings of this study have been deposited in the NCBI SRA database with the primary accession code PRJNA1453921.

