## Supplementary Tables and Figures for "Early transcriptional signature in chikungunya predicts chronic arthralgia and reveals deficient antiviral response"

Supplementary Material

### Supplementary Table 4: Log2 fold change and standard error of differentially expressed genes in chronic chikungunya, recovered chikungunya and control groups. The standard error was not corrected for multiple comparisons. The comparison recovered versus control for *NAV3* and *PTGER3* was not conducted due to low counts (less than 1 fragment per million in less than 10 samples)

| **Gene** | **log_2_FC - CC vs RC** | **SE - CC vs RC** | **FDR - CC vs RC** | **log_2_FC - CC vs Control** | **SE – CC vs Control** | **FDR – CC vs Control** | **log_2_FC - RC vs Control** | **SE - RC vs Control** | **FDR - RC vs Control** |
| --- | --- | --- | --- | --- | --- | --- | --- | --- | --- |
| *TNFRSF18* | 1.3 | 0.4 | 0.048 | 0.12 | 0.42 | 0.789 | -0.93 | 0.55 | 0.068 |
| *PTGER3* | 1.23 | 0.37 | 0.034 | 2.04 | 0.36 | <0.0001 | - | - | - |
| *ACKR3* | -1.69 | 0.42 | 0.018 | 0.16 | 0.38 | 0.676 | 2.14 | 0.49 | <0.0001 |
| *IKZF2* | 1.32 | 0.38 | 0.028 | -0.69 | 0.34 | 0.042 | -2.04 | 0.34 | <0.0001 |
| *TMEM176A* | -2.39 | 0.55 | 0.001 | -0.27 | 0.46 | 0.546 | 1.38 | 0.84 | 0.024 |
| *TMEM176B* | -2.47 | 0.64 | 0.003 | -1.05 | 0.53 | 0.031 | 0.55 | 0.74 | 0.284 |
| *NCS1* | 1.11 | 0.3 | 0.023 | -0.48 | 0.49 | 0.295 | -1.18 | 0.87 | 0.072 |
| *NAV3* | 2.32 | 0.72 | 0.034 | 3.51 | 0.79 | <0.0001 | - | - | - |
| *ST8SIA1* | 1.75 | 0.52 | 0.036 | -1.57 | 0.43 | <0.0001 | -3.42 | 0.33 | <0.0001 |
| *OLIG1* | -1.17 | 0.32 | 0.024 | -0.54 | 0.34 | 0.118 | 0.51 | 0.43 | 0.243 |

### Supplementary Table 5: Characteristics of the patients included in the qRT-PCR analyses.

| **Characteristic** | **Chronic** | **Recovered** | **p-value** |
| --- | --- | --- | --- |
|  | **N = 12** | **N = 12** |  |
| **Age - years, median (IQR)** | 45 (32 to 51) | 23 (17 to 35) | 0.04 |
| **Sex - Female** | 11 (92) | 3 (25) | <0.01 |
| **Symptom onset - time (days), median (IQR)** | 2.00 (1.00 to 5.50) | 1.00 (1.00 to 2.50) | 0.3 |
| **Hypertension** | 4 (33) | 0 (0) | 0.09 |
| **Diabetes** | 2 (17) | 1 (8.3) | >0.9 |
| **Arthalgia** | 12 (100) | 12 (100) |  |
| **Maculopapular Cutaneous Rash** | 4 (33) | 4 (33) | >0.9 |
| **Edema** | 9 (75) | 2 (17) | <0.01 |
| **Pruritus** | 3 (25) | 3 (25) | >0.9 |
| **Dizziness** | 11 (92) | 8 (67) | 0.3 |
| **Fever** | 12 (100) | 12 (100) |  |
| **Headache** | 10 (83) | 12 (100) | 0.5 |
| **Myalgia** | 11 (92) | 10 (83) | >0.9 |
| **Retro-orbital pain** | 8 (67) | 8 (67) | >0.9 |
| **Arthralgia duration (days), median (IQR)** | 725 (719 to 942) | 9 (2 to 18) | <0.01 |
| **qRT-PCR (Cycle threshold), median (IQR)** | 28.4 (26.0 to 32.2)* | 27.3 (25.0 to 30.0) | 0.4 |

IQR = Interquartile Range | qRT= real-time reverse transcription-PCR. P-value comparisons between chronic and recovered patients using the Wilcoxon rank sum test or Pearson’s Chi-squared test. Numbers are N(%) or otherwise specified. * One sample positive only in IgM.

Three patients (2 recovered and 1 chronic) were not included in the RNASeq analysi

### Supplementary Table 6. qRT-PCR analysis of pre-selected genes from RNASeq.

| **Gene** | **ΔCt - RC** | **SD** | **ΔCt - CC** | **SD** | **ΔΔCt** | **Fold Change** | **Log_2_FC** | **T test p-value** | **Wilcoxon p-value** |
| --- | --- | --- | --- | --- | --- | --- | --- | --- | --- |
| **Normalized by GAPDH** | | | | | | | | | |
| *ACKR3* | 5.940 | 2.622 | 7.184 | 1.658 | 1.244 | 0.422 | -1.244 | 0.181 | 0.291 |
| *IKZF2* | 10.399 | 1.738 | 8.221 | 2.060 | -2.178 | 4.525 | 2.178 | 0.024 | 0.041 |
| *CD45* | 0.250 | 2.267 | -0.003 | 1.513 | -0.253 | 1.192 | 0.253 | 0.751 | 0.551 |
| *TMEM176a* | 2.589 | 2.810 | 3.981 | 2.458 | 1.392 | 0.381 | -1.392 | 0.210 | 0.143 |
| *TMEM176b* | 3.158 | 2.522 | 4.723 | 1.759 | 1.566 | 0.338 | -1.566 | 0.093 | 0.101 |
| **Normalized by GAPDH and CD45** | | | | | | | | | |
| *ACKR3* | 5.815 | 1.889 | 7.186 | 1.378 | 1.371 | 0.387 | -1.371 | 0.056 | 0.046 |
| *IKZF2* | 9.865 | 1.240 | 8.213 | 1.480 | -1.652 | 3.142 | 1.652 | 0.017 | 0.020 |
| *TMEM176a* | 2.464 | 2.413 | 3.983 | 2.274 | 1.518 | 0.349 | -1.518 | 0.127 | 0.101 |
| *TMEM176b* | 3.033 | 1.994 | 4.725 | 1.759 | 1.693 | 0.309 | -1.693 | 0.038 | 0.039 |

### Supplementary Table 7: Baseline characteristics of patients with chronic chikungunya and recovered individuals.

| ***Characteristic*** | ***Chronic,***  ***N = 4*** | ***Recovered,***  ***N = 3*** | ***p-value*** |
| --- | --- | --- | --- |
| *Age - years, median (IQR)* | 45 (27, 62) | 18 (14,34) | 0.11 |
| *Sex - Female, number (%)* | 2 (50%) | 2 (75%) | 0.43 |
| *Time from symptom onset to assessment, days*  *(IQR)* | 4.5 (0.75, 59.5) | 5 (2, 5) | 0.86 |
| *Arthralgia* | 4 (100%) | - | - |
| ***Symptoms*** |  |  |  |
| *Maculopapular Cutaneous Rash* | 2 (50%) | 0 (0%) | 0.43 |
| *Articular edema* | 1 (25%) | 0 (0%) | >0.9 |
| *Fever* | 4 (100%) | 3 (75%) | >0.9 |
| *Headache* | 2 (50%) | 1 (25%) | >0.9 |

IQR = Interquartile Range | qRT= real-time reverse transcription-PCR

### Supplementary Table 8: Characteristics of Luminex patients

| **Characteristic** | **Chronic, N=27** | **Recovered, N=11** | **p-value** |
| --- | --- | --- | --- |
| Age - years, median (IQR) | 42 (29 to 54) | 23 (15 to 43) | 0.046 |
| Sex - Female | 19 (70) | 4 (36) | 0.073 |
| Symptom onset - time | 1.00 (1.00 to 3.00) | 1.00 (1.00 to 3.00) | >0.9 |
| Hypertension | 5 (19) | 0 (0) | 0.3 |
| Diabetes | 4 (15) | 1 (9.1) | >0.9 |
| Arthalgia | 27 (100) | 11 (100) |  |
| Maculopapular Cutaneous Rash | 11 (41) | 3 (27) | 0.5 |
| Edema | 21 (78) | 1 (9.1) | <0.001 |
| Pruritus | 12 (44) | 3 (27) | 0.5 |
| Dizziness | 19 (70) | 9 (82) | 0.7 |
| Fever | 27 (100) | 11 (100) |  |
| Headache | 24 (89) | 11 (100) | 0.5 |
| Myalgia | 25 (93) | 9 (82) | 0.6 |
| Retro-orbital pain | 17 (63) | 8 (73) | 0.7 |
| Arthralgia duration (days), median (IQR) | 720 (353 to 948) | 11 (5 to 21) | <0.001 |
| qRT-PCR (Cycle threshold) | 26.4 (21.6 to 29.8) | 28.4 (24.6 to 32.2) | 0.5 |

IQR = Interquartile Range | qRT= real-time reverse transcription-PCR. P-value comparisons between chronic and recovered patients using the Wilcoxon rank sum test or Pearson’s Chi-squared test. Numbers are N(%) or otherwise specified

### Supplementary Table 9: Markers evaluated through Luminex using Wilcoxon rank sum p-value and Benjamini-Hochberg adjustment.

|  | **p-value** | **adjusted p-value (BH)** |
| --- | --- | --- |
| EGF | 0.46 | 0.75 |
| Eotaxin | 0.63 | 0.81 |
| G-CSF | 0.36 | 0.69 |
| GM-CSF | 0.38 | 0.69 |
| IFN-α2 | 0.37 | 0.69 |
| IFN-γ | 0.07 | 0.34 |
| IL-10 | 0.08 | 0.34 |
| IL-12p40 | 0.33 | 0.69 |
| IL-12p70 | 0.02 | 0.2 |
| IL-13 | 0.13 | 0.43 |
| IL-15 | 0.58 | 0.77 |
| IL-17A | 0.08 | 0.35 |
| IL-1RA | 0.12 | 0.43 |
| IL-1α | 0.33 | 0.69 |
| IL-1β | 0.07 | 0.34 |
| IL-2 | 0.36 | 0.69 |
| IL-3 | 0.29 | 0.69 |
| IL-4 | 0.5 | 0.75 |
| IL-5 | 0.33 | 0.69 |
| IL-6 | 0.01 | 0.14 |
| IL-7 | 0.01 | 0.14 |
| IL-8 | 0.01 | 0.14 |
| IP-10 | 0.55 | 0.75 |
| MCP-1 | 0.16 | 0.48 |
| MIP-1α | 0 | 0.14 |
| MIP-1β | 0.06 | 0.34 |
| SDF-1α+β | 0.78 | 0.82 |
| TGF-β1 | 0.68 | 0.81 |
| TNF-α | 0.55 | 0.75 |
| TNF-β | 0.06 | 0.34 |
| VEGF | 0.19 | 0.52 |
| sCD30 | 0.14 | 0.43 |
| sEGFR | 1 | 1 |
| sIL-1RI | 0.65 | 0.81 |
| sIL-1RII | 0.66 | 0.81 |
| sIL-2Rα | 0.52 | 0.75 |
| sIL-4R | 0.55 | 0.75 |
| sIL-6R | 0.95 | 0.97 |
| sRAGE | 0.46 | 0.75 |
| sTNFRI | 0.2 | 0.52 |
| sTNFRII | 0.73 | 0.82 |
| sVEGFR1 | 0.77 | 0.82 |
| sVEGFR2 | 0.47 | 0.75 |
| sVEGFR3 | 0.71 | 0.82 |
| sgp130 | 0.77 | 0.82 |

### Supplementary Figure 1. Volcano plot depicting differentially expressed genes in the comparison between chronic and recovered chikungunya patients

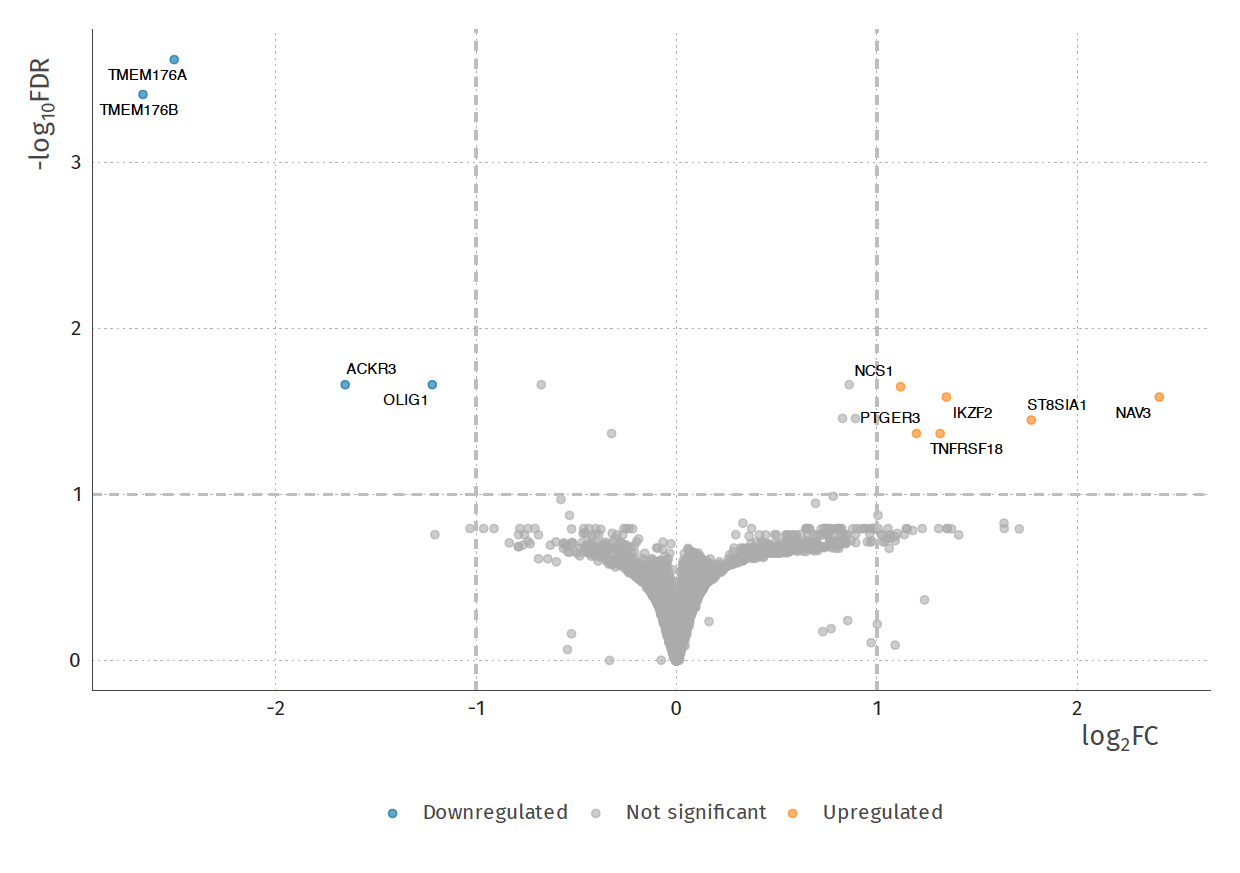

### Supplementary Figure 2: Flow cytometry gate scheme.

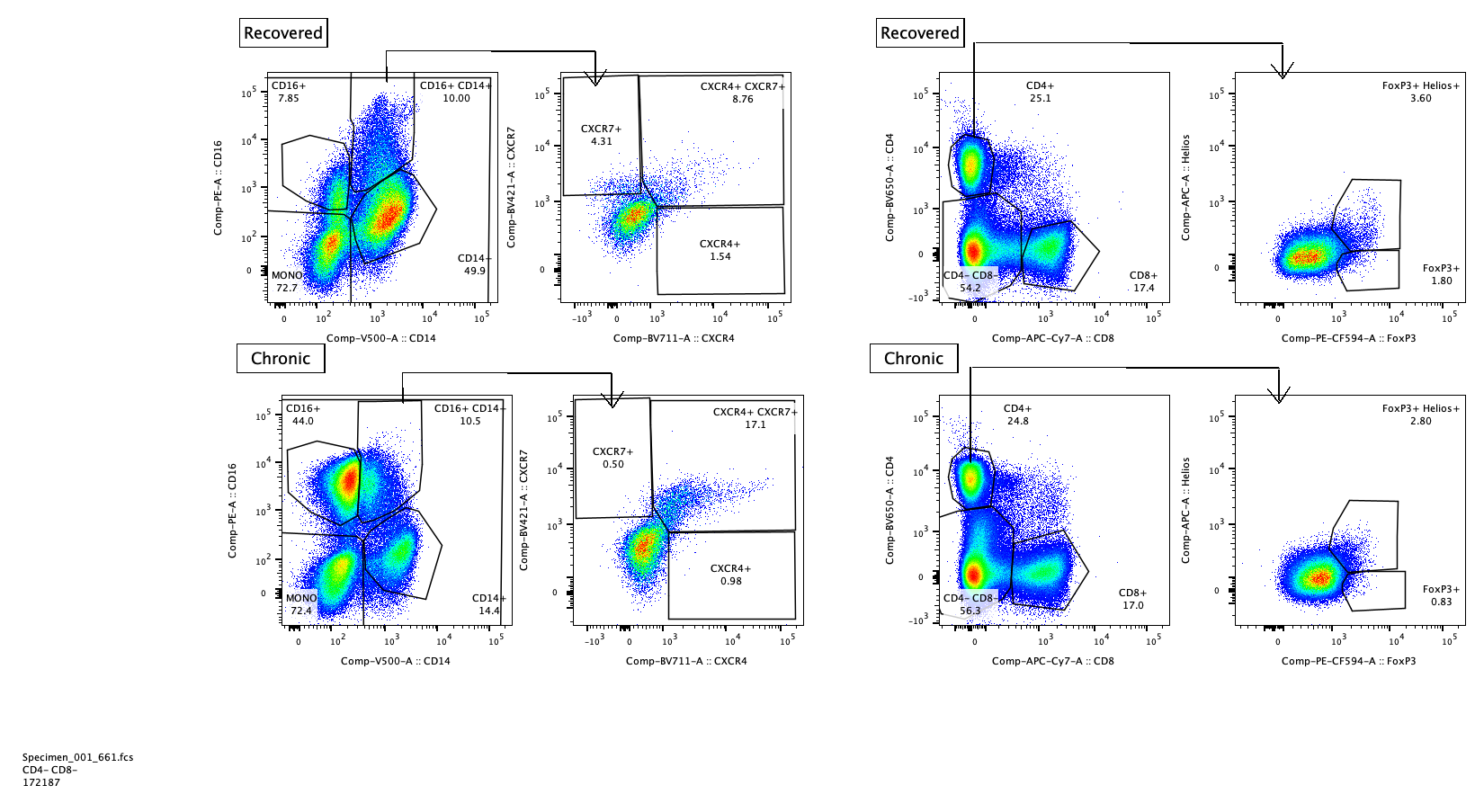

### Supplementary Figure 3: Volcano plot depicting differentially expressed genes in the comparison between chronic and recovered chikungunya male patients

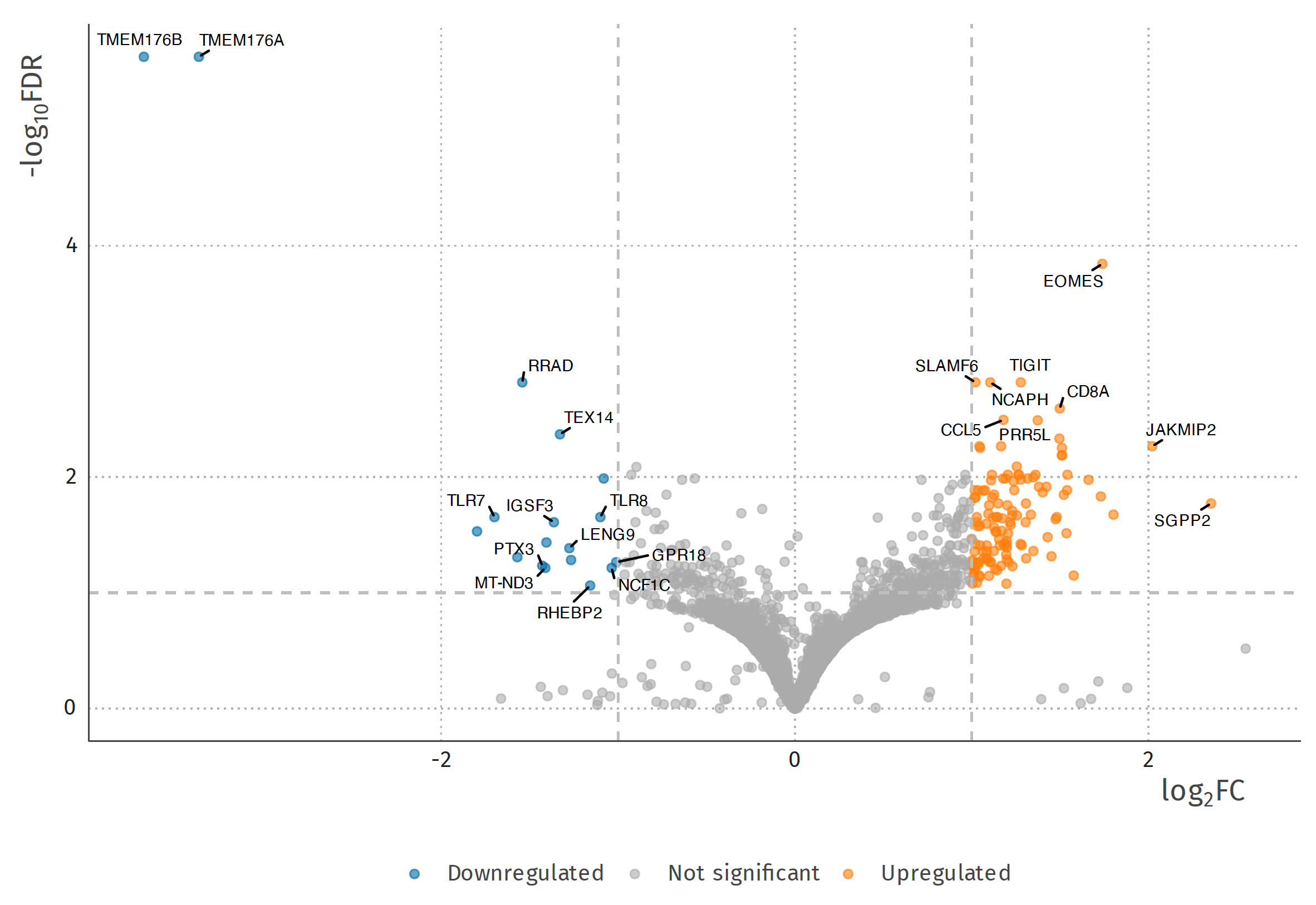

### Supplementary Figure 4: Volcano plot depicting differentially expressed genes in the comparison between chronic and recovered chikungunya female patients

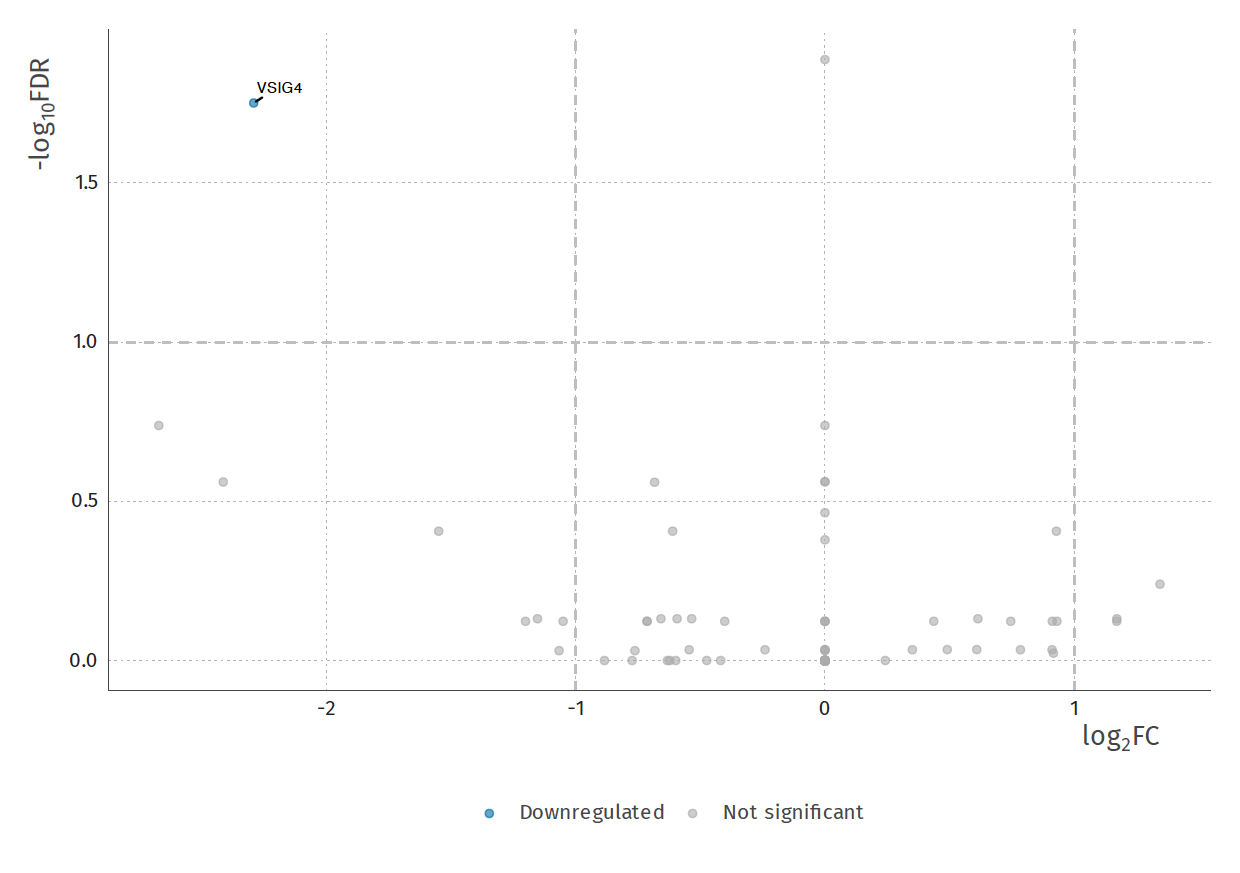

### Supplementary Figure 5: Dot plot of the expression comparing chronic and recovered chikungunya patients in the Luminex experiment

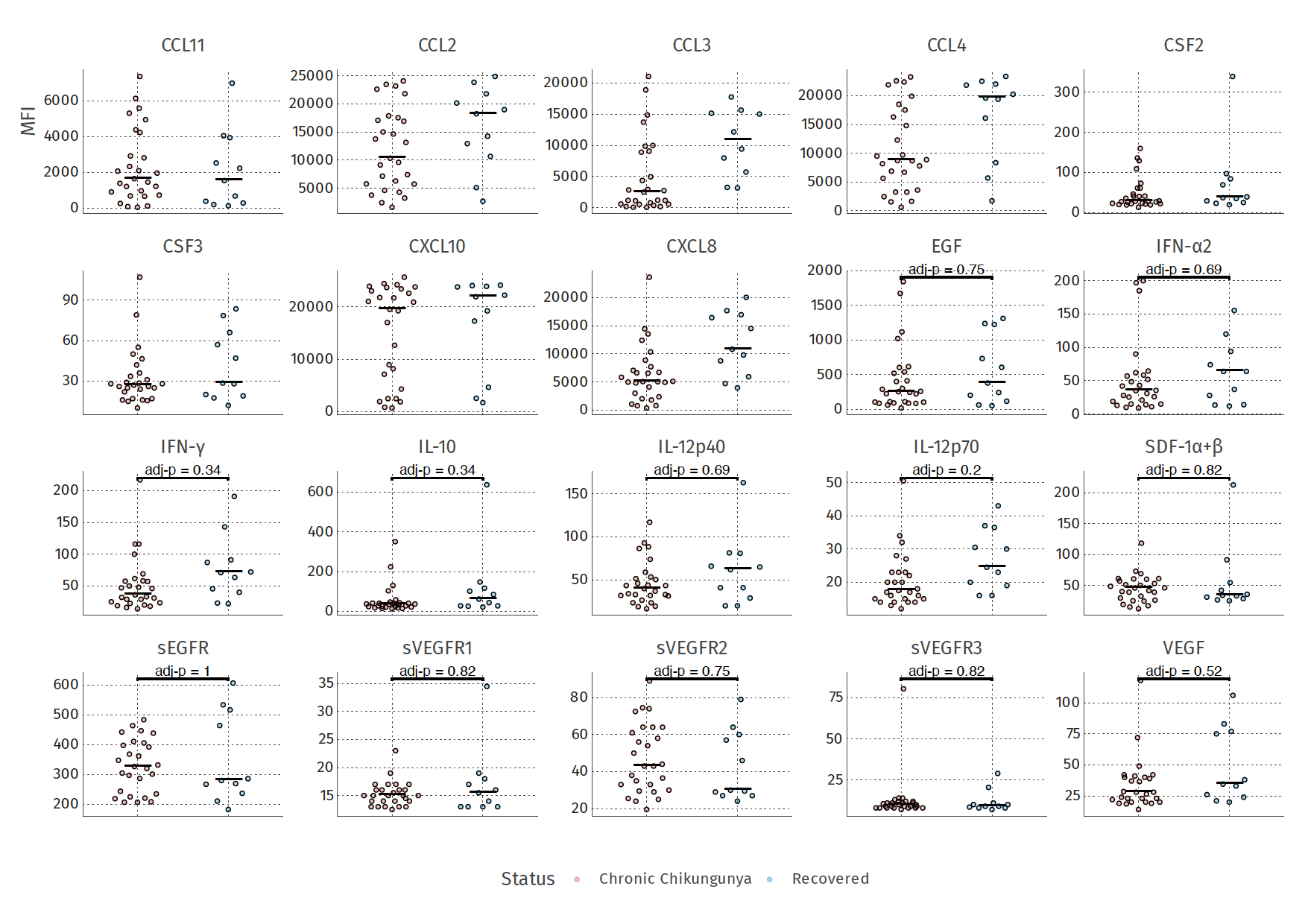

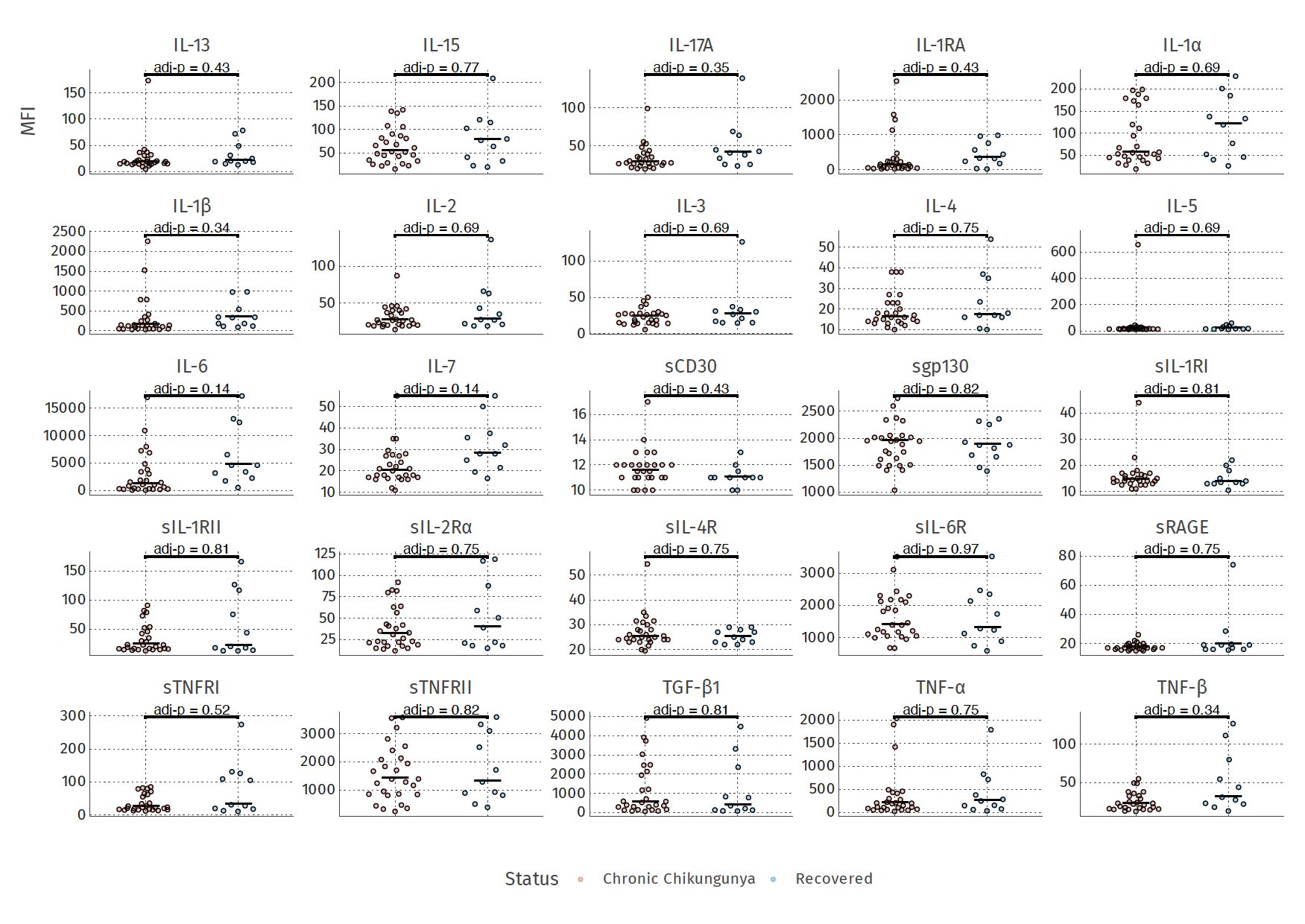
